# Latent tuberculosis infection workflows in four primary healthcare systems in the United States

**DOI:** 10.64898/2026.06.12.26355200

**Authors:** Julie Espey, Julie Venci, Paul Wada, Yoseph Sorri, Amy Tang, Beatrice Francis, Jacek Skarbinski, Masahiro Narita, Priya B. Shete, Kathryn Winglee, the Tuberculosis Epidemiologic Studies Consortium

**Affiliations:** Division of Tuberculosis Elimination, National Center for HIV, Viral Hepatitis, STD, and TB Prevention, Centers for Disease Control and Prevention, Atlanta, GA, USA; Denver Health and Hospital Authority, Denver, CO, USA; The Permanente Medical Group, Kaiser Permanente Northern California, Pleasanton, CA, 94588; Public Health-Seattle and King County, Seattle, WA, USA; North East Medical Services, San Francisco, CA, USA; Public Health Institute at Denver Health, Denver, CO, USA; University of California San Francisco, San Francisco, CA, USA

## Abstract

**Background:** Latent tuberculosis - infection (LTBI) reactivation accounts for most tuberculosis (TB) cases in the United States (U.S.). Expanding primary care LTBI services can improve detection and treatment, but real-world implementation is not well described.

**Objective:** Describe how LTBI screening and treatment workflows are operationalized into practice in diverse U.S. primary care health systems.

**Design:** Descriptive, multi-site study.

**Setting:** Four U.S. primary care health systems participating in the U.S. Centers for Disease Control and Prevention (CDC) Tuberculosis Epidemiologic Studies Consortium III (TBESC-III, 2020-2022).

**Patients:** Patients visiting any study site (N = 3,522,077).

**Measurements:** Electronic medical record (EMR) data on patient demographics, TB test type, and results. Using narrative reporting and study documentation, LTBI care workflows were summarized by site into six domains: patient registration, TB testing evaluation, TB testing, TB disease assessment, LTBI diagnosis, and LTBI treatment.

**Results:** Sites implemented recommended practices for testing and treatment: interferon-gamma release assays were the predominant test, and short-course rifamycin-based regimens were the primary treatment. Follow-up care practices and documentation of treatment outcomes varied. Country of birth was not consistently recorded at all sites, limiting TB screening capability based on nativity. Physician training, EMR decision-support tools, and patient education resources differed in scope and availability.

**Limitations:** Descriptive design using self-reported workflows; may not generalize to all primary care settings or reflect physician-level variations.

**Conclusion:** We provide a descriptive snapshot showing how LTBI care workflows varied in approach and consistency at four primary care systems with integrated LTBI services. These real-world examples may serve as templates for U.S. primary care systems seeking to expand TB testing and treatment.

**Primary Funding Source:** U.S. Centers for Disease Control and Prevention (TBESC-III).

## Background

In the United States, most tuberculosis (TB) cases arise from reactivation of latent tuberculosis infection (LTBI) rather than recent transmission (1,2), and the Centers for Disease Control and Prevention (CDC) estimates up to 13 million individuals are living in the United States with LTBI (3). This reactivation risk falls disproportionately on non-U.S.– born persons, among whom LTBI prevalence (15.9–20.5%) is nearly 10-fold higher than U.S.-born individuals (1.5–2.8%) (3). The observed disparity underlies a substantial public health burden for the United States where approximately 7–10 thousand TB cases have been reported annually between 2020 and 2024, with 470–602 deaths per year attributed to TB disease (4). Even with completed therapy, over half of TB survivors may experience long-term pulmonary, cardiovascular, or other sequelae (5), resulting in excess morbidity, mortality, and disability (5,6). Further, case counts have been rising since 2021 (4), making effective LTBI detection and treatment all the more urgent for advancing U.S. TB elimination efforts (7).

Decentralizing LTBI care from public health and expanding services into primary care is a key strategy to improve detection and treatment in the United States (3,8–10). Because LTBI is an asymptomatic condition, affected individuals are unlikely to seek care at public health TB clinics where TB-related health services traditionally are concentrated (11– 14). Integrating LTBI care into routine primary care may expedite diagnosis and treatment of both LTBI and TB disease – particularly among non-U.S.–born individuals from TB-endemic countries (8,13,14). In the United States, non-U.S.–born persons represent 76% of all TB cases, underscoring the importance of TB healthcare for this population to accelerate efforts to eliminate TB (12).

The U.S. CDC and the U.S. Preventive Services Task Force (USPSTF)provide clinical recommendations for LTBI care (8), but there is limited literature detailing how recommendations are implemented in real-world practice across diverse clinical settings (15). To address this gap, we described LTBI screening and treatment practices at four U.S. primary care sites during the 2020–2022 period of CDC’s third Tuberculosis Epidemiologic Studies Consortium (TBESC-III) (16). This study characterizes LTBI care workflows, care approaches, and operational challenges across the Consortium to provide a descriptive snapshot of routine LTBI care.

## Methods

### Study design and data collection

In 2021, TBESC-III funded four sites to examine and enhance LTBI screening, diagnosis, and treatment practices within primary care settings (17) in the United States (16,18). Each site was required to have an electronic medical record (EMR) system and serve at least 10,000 non-U.S.–born persons at increased risk for TB infection, based on high rates of TB disease among their expatriates currently living in the United States (≥10 cases per 100,000 persons) (19). Clinical sites designated, at minimum, a one-year study period, with study start dates varying across sites (Table 1). This period was selected to best describe contemporary practices at the start of the study and capture EMR data on patient demographics, visits, diagnostics, and treatment history.

**Table 1.**
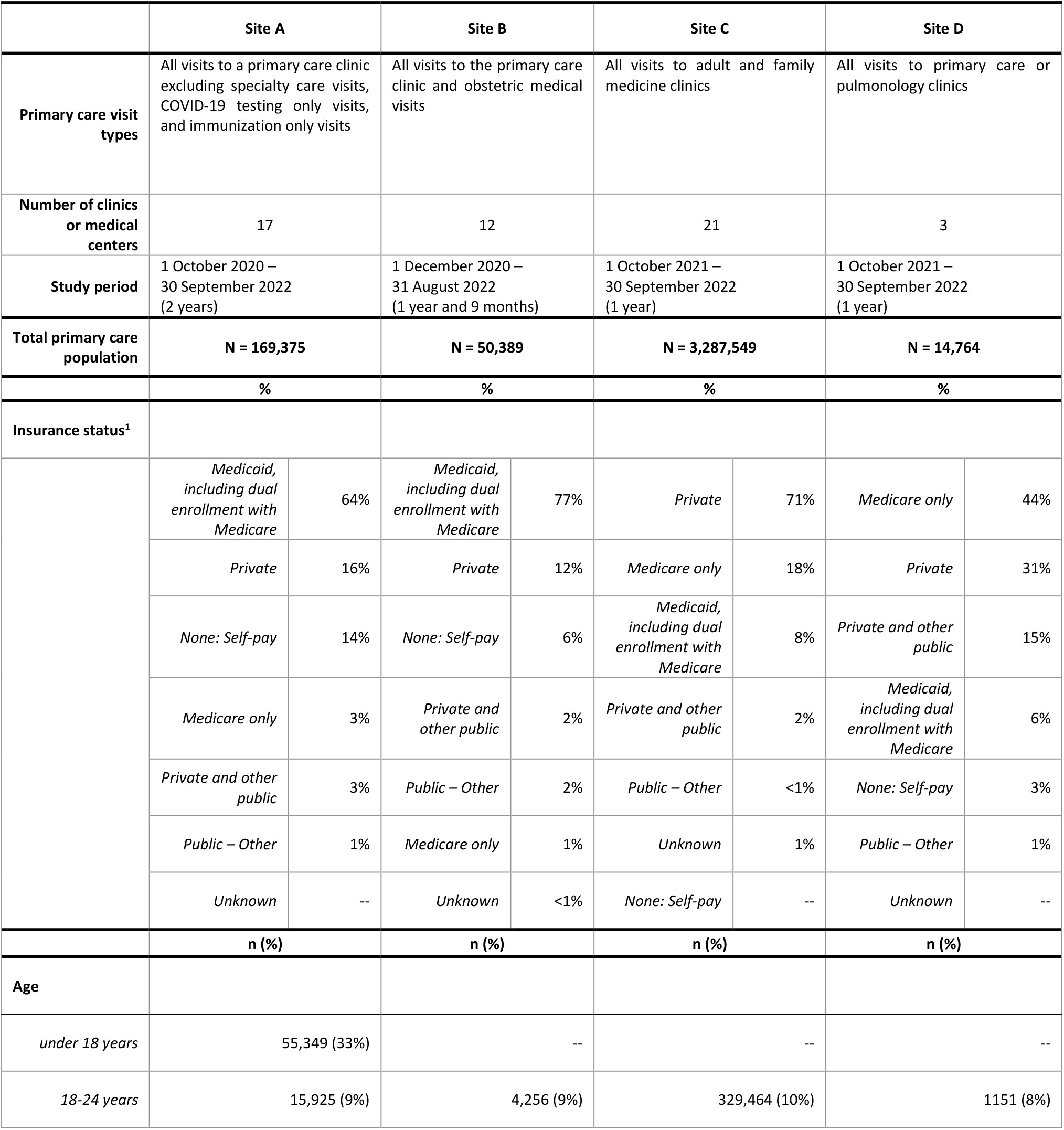
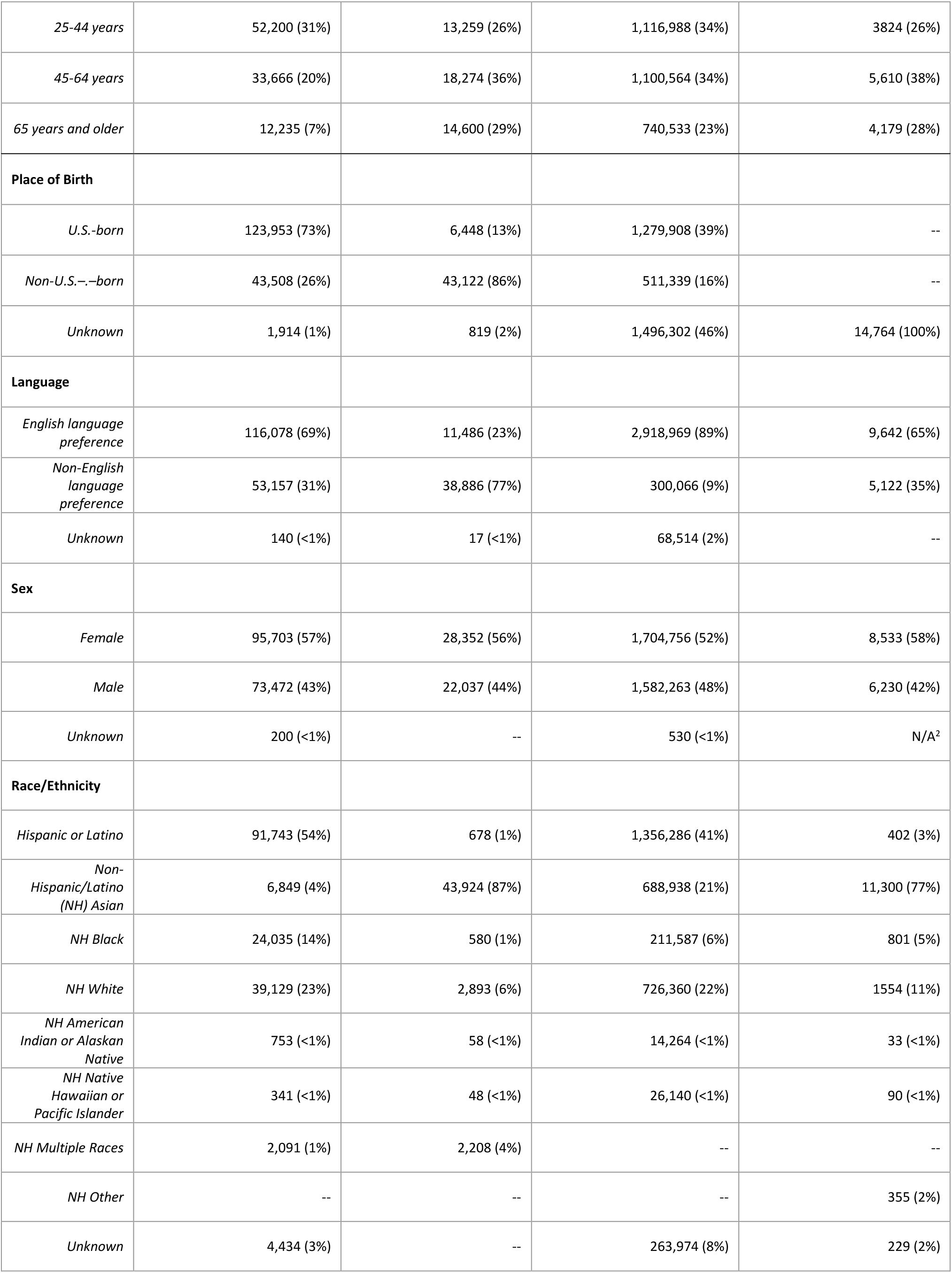
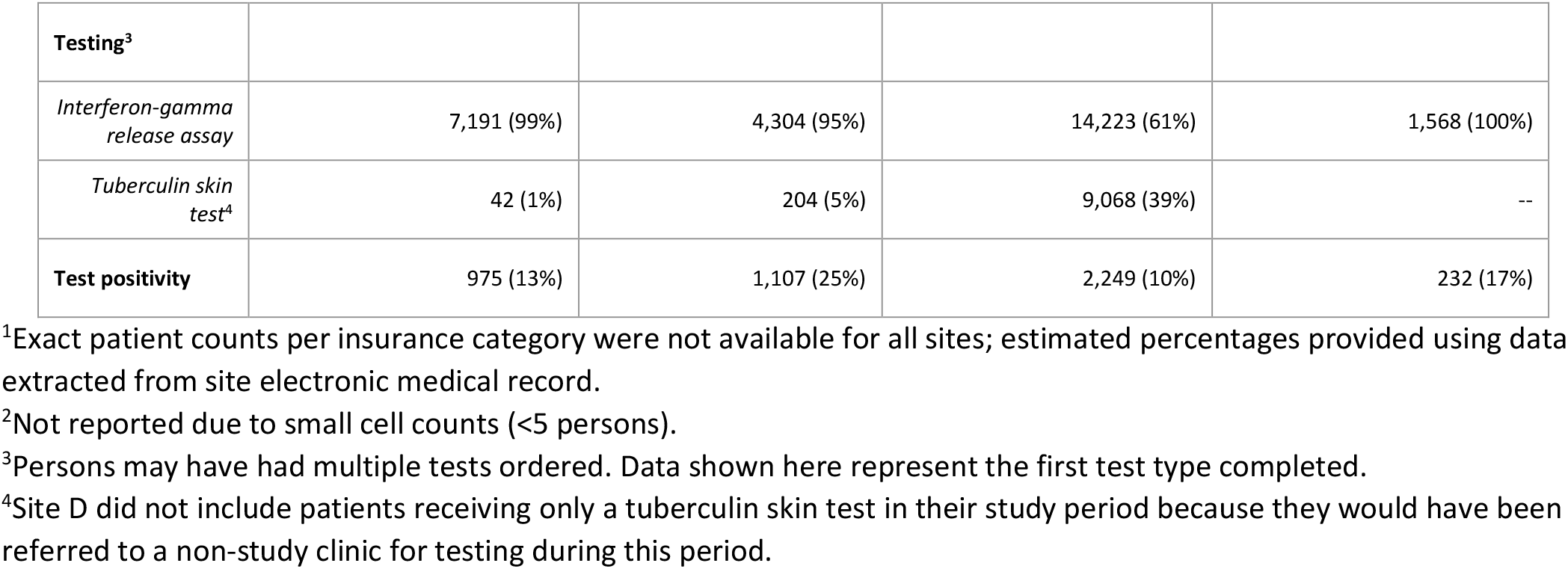
Key characteristics of Tuberculosis Epidemiologic Studies Consortium III primary care settings and patients, 2020––2022.

|  | Site A |  | Site B |  | Site C |  | Site D |  |
| --- | --- | --- | --- | --- | --- | --- | --- | --- |
| **Primary care visit types** | All visits to a primary care clinic excluding specialty care visits, COVID-19 testing only visits, and immunization only visits |  | All visits to the primary care clinic and obstetric medical visits |  | All visits to adult and family medicine clinics |  | All visits to primary care or pulmonology clinics |  |
| **Number of clinics or medical centers** | 17 |  | 12 |  | 21 |  | 3 |  |
| **Study period** | 1 October 2020 – 30 September 2022 (2 years) |  | 1 December 2020 – 31 August 2022 (1 year and 9 months) |  | 1 October 2021 – 30 September 2022 (1 year) |  | 1 October 2021 – 30 September 2022 (1 year) |  |
| **Total primary care population** | **N = 169,375** |  | **N = 50,389** |  | **N = 3,287,549** |  | **N = 14,764** |  |
|  | **%** |  | **%** |  | **%** |  | **%** |  |
| **Insurance status<sup>1</sup>** |  |  |  |  |  |  |  |  |
|  | *Medicaid, including dual enrollment with Medicare* | 64% | *Medicaid, including dual enrollment with Medicare* | 77% | *Private* | 71% | *Medicare only* | 44% |
|  | *Private* | 16% | *Private* | 12% | *Medicare only* | 18% | *Private* | 31% |
|  | *None: Self-pay* | 14% | *None: Self-pay* | 6% | *Medicaid, including dual enrollment with Medicare* | 8% | *Private and other public* | 15% |
|  | *Medicare only* | 3% | *Private and other public* | 2% | *Private and other public* | 2% | *Medicaid, including dual enrollment with Medicare* | 6% |
|  | *Private and other public* | 3% | *Public – Other* | 2% | *Public – Other* | <1% | *None: Self-pay* | 3% |
|  | *Public – Other* | 1% | *Medicare only* | 1% | *Unknown* | 1% | *Public – Other* | 1% |
|  | *Unknown* | -- | *Unknown* | <1% | *None: Self-pay* | -- | *Unknown* | -- |
|  | **n (%)** |  | **n (%)** |  | **n (%)** |  | **n (%)** |  |
| **Age** |  |  |  |  |  |  |  |  |
| *under 18 years* | 55,349 (33%) |  | -- |  | -- |  | -- |  |
| *18-24 years* | 15,925 (9%) |  | 4,256 (9%) |  | 329,464 (10%) |  | 1151 (8%) |  |

|  |  |  |  |  |
| --- | --- | --- | --- | --- |
| *25-44 years* | 52,200 (31%) | 13,259 (26%) | 1,116,988 (34%) | 3824 (26%) |
| *45-64 years* | 33,666 (20%) | 18,274 (36%) | 1,100,564 (34%) | 5,610 (38%) |
| *65 years and older* | 12,235 (7%) | 14,600 (29%) | 740,533 (23%) | 4,179 (28%) |
| **Place of Birth** |  |  |  |  |
| *U.S.-born* | 123,953 (73%) | 6,448 (13%) | 1,279,908 (39%) | -- |
| *Non-U.S.-born* | 43,508 (26%) | 43,122 (86%) | 511,339 (16%) | -- |
| *Unknown* | 1,914 (1%) | 819 (2%) | 1,496,302 (46%) | 14,764 (100%) |
| **Language** |  |  |  |  |
| *English language preference* | 116,078 (69%) | 11,486 (23%) | 2,918,969 (89%) | 9,642 (65%) |
| *Non-English language preference* | 53,157 (31%) | 38,886 (77%) | 300,066 (9%) | 5,122 (35%) |
| *Unknown* | 140 (<1%) | 17 (<1%) | 68,514 (2%) | -- |
| **Sex** |  |  |  |  |
| *Female* | 95,703 (57%) | 28,352 (56%) | 1,704,756 (52%) | 8,533 (58%) |
| *Male* | 73,472 (43%) | 22,037 (44%) | 1,582,263 (48%) | 6,230 (42%) |
| *Unknown* | 200 (<1%) | -- | 530 (<1%) | N/A<sup>2</sup> |
| **Race/Ethnicity** |  |  |  |  |
| *Hispanic or Latino* | 91,743 (54%) | 678 (1%) | 1,356,286 (41%) | 402 (3%) |
| *Non-Hispanic/Latino (NH) Asian* | 6,849 (4%) | 43,924 (87%) | 688,938 (21%) | 11,300 (77%) |
| *NH Black* | 24,035 (14%) | 580 (1%) | 211,587 (6%) | 801 (5%) |
| *NH White* | 39,129 (23%) | 2,893 (6%) | 726,360 (22%) | 1554 (11%) |
| *NH American Indian or Alaskan Native* | 753 (<1%) | 58 (<1%) | 14,264 (<1%) | 33 (<1%) |
| *NH Native Hawaiian or Pacific Islander* | 341 (<1%) | 48 (<1%) | 26,140 (<1%) | 90 (<1%) |
| *NH Multiple Races* | 2,091 (1%) | 2,208 (4%) | -- | -- |
| *NH Other* | -- | -- | -- | 355 (2%) |
| *Unknown* | 4,434 (3%) | -- | 263,974 (8%) | 229 (2%) |

| Testing<sup>3</sup> |  |  |  |  |
| --- | --- | --- | --- | --- |
| *Interferon-gamma release assay* | 7,191 (99%) | 4,304 (95%) | 14,223 (61%) | 1,568 (100%) |
| *Tuberculin skin test<sup>4</sup>* | 42 (1%) | 204 (5%) | 9,068 (39%) | -- |
| **Test positivity** | 975 (13%) | 1,107 (25%) | 2,249 (10%) | 232 (17%) |

<sup>1</sup>Exact patient counts per insurance category were not available for all sites; estimated percentages provided using data extracted from site electronic medical record.
<sup>2</sup>Not reported due to small cell counts (<5 persons).
<sup>3</sup>Persons may have had multiple tests ordered. Data shown here represent the first test type completed.
<sup>4</sup>Site D did not include patients receiving only a tuberculin skin test in their study period because they would have been referred to a non-study clinic for testing during this period.

Each site defined eligible primary care visits and age minimums during the study period (2020–2022) (Table 1). Visit types included both telehealth and in-person visits, with Site B including all 28-week prenatal visits for routine, universal TB screening of pregnant persons and Site D including only prenatal visits where risk-based screening was conducted. Site A included persons aged 2 years and older while Sites B, C, and D included only adults.

In December 2023, sites extracted data on patients in the study period from their EMR databases to generate summary statistics on demographic information, including age, sex, and race/ethnicity. Sites reported information on patients receiving a TB test, including the test type (i.e., interferon gamma release assay [IGRA] or tuberculin skin test [TST]) administered and test results. While TBESC-III collected LTBI treatment prescription and refill data for the non-U.S.–born population (18), this study describes all persons visiting a study site during the baseline study period, and treatment data were not available for this broader population.

### LTBI care model categories

At the conclusion of the study period, study staff provided documentation and retrospective information on typical LTBI care practices, including the healthcare staff involved in processes, aggregated patient demographics and testing outcome data, and challenges noted by healthcare practitioners throughout the LTBI workflow. Additionally, they reported information on available LTBI resources for primary care physicians (PCPs) and patient guidance. We consulted site staff and documentation on clinical activities to organize LTBI care processes into six sequential categories comprising a general workflow: patient registration, TB testing evaluation, TB testing, TB assessment, LTBI diagnosis, and LTBI treatment. Finally, we synthesized and visually depicted the typical flow of events from one category of the LTBI workflow to the next for each site for the Consortium (Figure 1) and within each site (Supplemental Figures 1–4).

**Figure 1.**
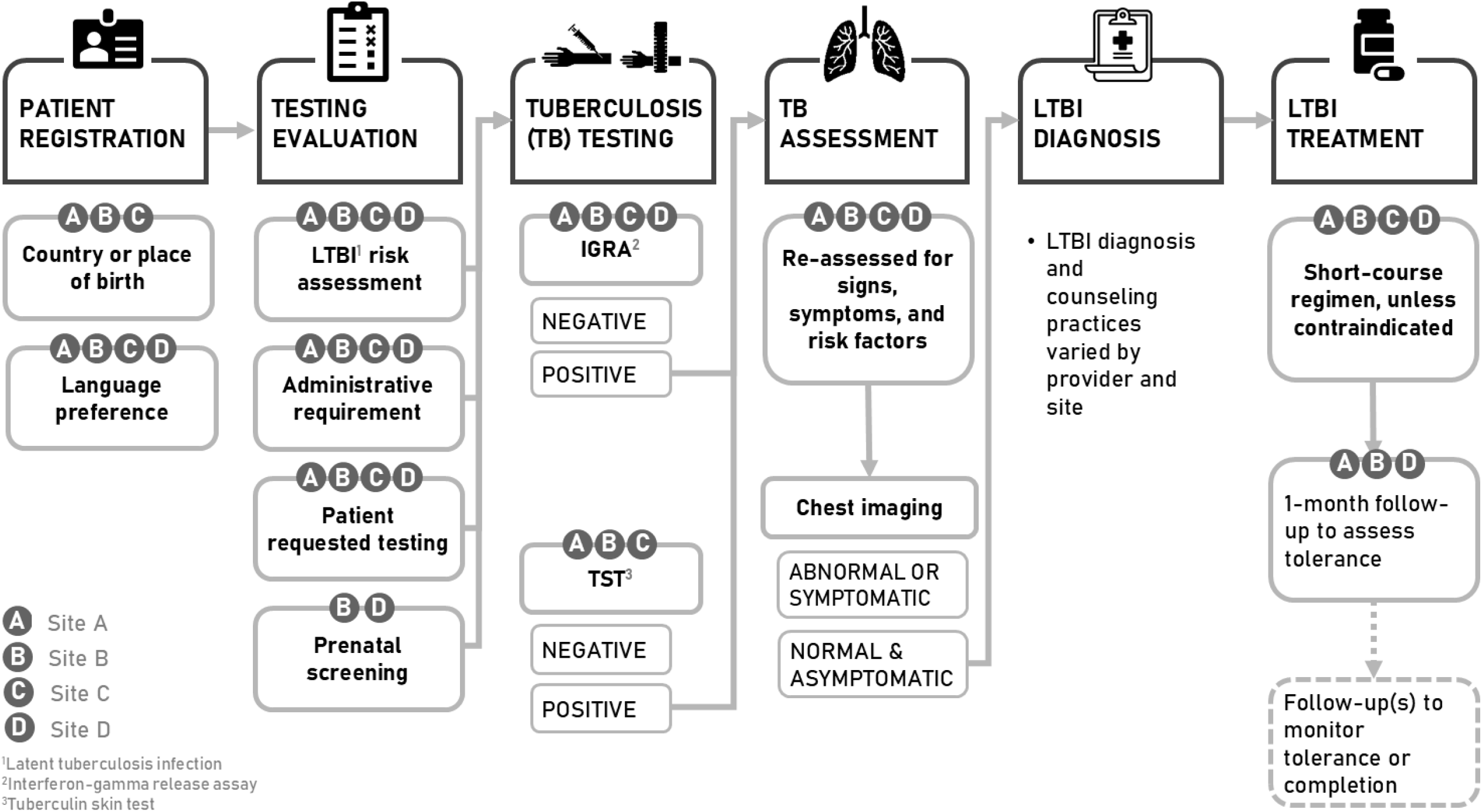
Summary of latent tuberculosis infection (LTBI) care workflows and clinical practices across four primary healthcare systems, Tuberculosis Epidemiologic Studies Consortium-III, 2020–2022.

### Ethical considerations

This study was reviewed by and conducted under the authority of the CDC. It was determined to not be human subjects research as the primary intent of the study was program evaluation and the study was conducted consistent with applicable federal law and CDC policy.

## Results

### Patient and Site Characteristics

Primary care populations ranged in size from 14,764 (Site D) to 3,287,549 patients (Site C), and study sites comprised at least three clinics or medical centers (3–21 clinics/centers) (Table 1). Only Site A included patients <18 years of age (33%) in their study population. Sites A and B had the largest share of patients using Medicaid either with or without dual Medicare enrollment (64% and 77%, respectively). Site C, the largest healthcare system spanning 21 centers, utilized an integrated membership insurance plan, and therefore had no uninsured patients and the largest proportion using private health insurance (71%). Site D skewed Medicare only (44%) followed by private coverage (31%). Among the three sites with country of birth data, there was a wide range in the proportion of patients born outside the United States (16%–86%). Site D used non-English language preference (35%) as a proxy for being non-U.S.–born. Sites A and C served mostly persons with Hispanic/Latino ethnicity (A: 54%, C: 41%). Sites B and D’s patient populations were overwhelmingly non-Hispanic Asian (B: 87%, D: 77%).

### LTBI Care Workflows

We summarized main similarities and differences between site LTBI care models in Figure 1 and reported site-specific care models as supplementary figures (Supplemental Figures 1–4).

#### 1. Patient Registration

Information on country of birth or language was collected and available to providers with varying degrees of completeness. Country of birth was collected and accessible to providers at three sites only – Sites A, B, and C – with completeness ranging from 54% to 99% (Table 1). Language preference, collected across all four sites, was more complete than nativity data, ranging from 98–100% completeness. Generally, demographic data were recorded either at patient registration events like visit check-ins and enrollment or re-enrollment for primary care services (Figure 1). Site D did not collect country of birth information during patient registration, but providers could record this information themselves in the EMR if disclosed by the patient. As a result, Site D generally relied on language preference as a proxy for country of birth during risk-based TB screening and reported 35% of their primary care population preferred the use of a non-English language.

Site C’s PCPs could update a patient’s primary written and spoken language information during a visit if this information was missing from earlier data collection points, but country of birth data entries lacked standardization and were not easily summarized with simple data queries. Site C also relied on language preference as a proxy for unknown country of birth.

#### 2. Testing Evaluation

Evaluations for TB test ordering were conducted by the PCP, who typically initiated a test order for one of four reasons: having an increased risk for TB infection (i.e., a positive LTBI risk assessment), an administrative requirement, patient-requested testing, or prenatal screening (Figure 1).

##### LTBI Risk Assessment

PCPs conducted LTBI risk assessments on an individual basis across sites using their clinical judgment paired with current healthcare guidance in their shared decision-making with the patient. Starting in May 2021, Site A had an EMR-based quality suggestion for providers that displayed at the bottom of a standard primary care note template if a patient was born in a high-risk country for TB and had not previously had an IGRA with a valid result. Site B had EMR-based prompts to complete annual TB risk assessments for two groups: (1) any patient with a prior negative or inconclusive TB test and (2) any patient born in a high-risk country with no prior TB testing. The second group required standardized assessment of country of birth to complete their risk assessments (88% known). All sites primarily ordered an IGRA over TST. Reasons for opting out of a risk assessment or test order by the provider or patients were not recorded systematically.

##### Administrative Requirement

The second common pathway for ordering a TB test was screening for administrative requirements, like testing for school or work. Here, providers ordered the test type requested, commonly a TST. Site C had an especially large volume of administrative testing with TSTs due to specific requirements from employers engaged in their insurance membership plans and neighboring universities (39%; Table 1).

##### Patient-requested Testing

Every site ordered tests for patients when requested. IGRA tests were the primary test type ordered in these situations. If close contact with a confirmed TB disease case was a factor, sites typically referred the patient to their local public health TB clinic for testing and further evaluation. Public health staff then conducted contact tracing and provided appropriate TB care for any signs of active disease. Site B had workflows in place for patients with a close TB contact whereby local public health clinics referred close contacts identified through tracing efforts to PCPs for repeat IGRA testing eight weeks post-exposure.

##### Prenatal Screening

Site B conducted routine TB screening at the 28-week prenatal visit, while Site D only conducted risk-based testing during prenatal services. Both sites B and D performed prenatal screening primarily using an IGRA test.

##### TB Testing

Tests were conducted at the primary care clinic and ordered by PCPs. Site A referred patients to the public health TB clinic if a TST was requested and not currently available, otherwise it was administered in the clinic by medical staff other than the PCP. In general, negative IGRA and TST results were assumed to be true negatives by all sites prompting no additional workup, though TB disease assessments and further evaluation still occurred at the discretion of the PCP. For those reporting a close TB contact, repeat IGRA tests were ordered 8 weeks after a negative test for all sites. Person with a positive TST test and history of Bacillus Calmette-Guerin vaccination had confirmatory IGRA tests ordered at all sites. Indeterminate, invalid, and borderline IGRA tests were repeated.

In the Consortium, 36,418 persons (1.0%) were tested for TB infection at least once, with an average test positivity of 16% (range 10–25%) among primary care patients (Table 1). On average, 89% (range 61–100%) were tested with an IGRA first. Site D did not use TSTs for TB screening during this period, and any patients requesting a TST were referred to a non-study clinic. Even without Site D, we saw a comparably high adoption of IGRAs (85%; range: 61–99%).

#### 3. TB Assessment

Patients with a negative test and no reason for further TB evaluation exited the LTBI workflow at this step. Patients with a positive IGRA or TST were re-assessed for signs, symptoms, and risk factors for TB disease. Chest imaging – with a chest x-ray (CXR) or computed tomography (CT) scan – was a critical part of this assessment to rule out pulmonary TB disease. Three sites used recent imaging pre-dating a positive TB test when available to rule out pulmonary TB. At Site A, asymptomatic patients at high risk of progression to TB disease (e.g., children under 5 years old, contacts of TB disease, persons with HIV or otherwise immunocompromised) required a CXR within 2 months.

Otherwise, imaging within 6 months was acceptable. Site B also accepted imaging within 6 months of a positive TB test if there were no new symptoms. Site D had a narrower time frame of 3 months. Sites A and B frequently referred patients with suspected TB disease to public health TB clinics for further evaluation, while Sites C and D conducted TB evaluation on-site utilizing smear microscopy, cultures, and nucleic acid amplification testing as additional diagnostics for suspected pulmonary TB disease. Procedures to follow up on incomplete TB disease assessments were minimal or absent across sites. Additionally, the outcomes of patients referred to public health TB clinics were often unknown or not easily accessible for the PCP.

#### 4. LTBI Diagnosis

In the absence of symptoms or other risk factors warranting further evaluation, patients with normal chest imaging results were diagnosed with LTBI by their PCP. LTBI counseling practices (e.g., diagnosis consultation and resource sharing) varied by provider and site, with PCPs providing primary counseling and additional resources like translated LTBI materials wherever possible.

#### 5. Treatment

In accordance with national guidelines, short-course rifamycin-based LTBI regimens (i.e., four months of rifampin, three months of rifampin plus isoniazid, or 12 weeks of rifapentine plus isoniazid) (20) were the primary therapy recommended at all sites, unless there was a medical contraindication or other side effects of concern. Treatment monitoring practices generally were not standardized or comprehensively documented. Most patients diagnosed with LTBI at Site A were referred to the public health TB clinic for treatment prescription and management; Site A’s providers had EMR sharing privileges with their local TB clinic. Site B provided one- or two-month medication supplies initially and then monitored patients at variable intervals from monthly to every-few-months. Site C’s treatment monitoring varied based on each provider’s workload and time constraints, as monthly evaluations could be challenging to sustain and were not standard practice. Site D provided standardized LTBI treatment guidelines to all PCPs, including specific recommendations for prescribing rifamycin-based regimens, and generally, all LTBI patients had at least one follow-up visit a month after treatment initiation to assess tolerance and side effects.

Subsequent follow-up visits varied by PCPs within and across sites and depended on physician case load and availability. LTBI treatment was sometimes forgone for patients with complex conditions. For example, patients of advanced age (e.g., ≥75 years), individuals with significant medication interactions, or those on hospice care may have been deemed inappropriate candidates for LTBI treatment by their PCPs.

### Primary care physician and patient resources across sites

Most sites had some PCP training or other resources aimed at improving LTBI knowledge and supporting clinical decision making. Site A had clinical care guidelines developed by the local public health TB clinic on an intranet site to provide guidance on how to screen, diagnose and treat LTBI. Sites A and B had PCP-facing EMR tools in place for LTBI care management. Only Site D had support from other primary care practitioners like medical assistants for LTBI care management and TB testing with a TST (Supplemental Figure 4). Sites B and C had available translation services or linguistically appropriate LTBI resources tailored to persons whose preferred language was Chinese and Spanish, respectively. Site D also had translation services available, but no related LTBI educational materials translated into non-English languages.

## Discussion

Our descriptive assessment examines how LTBI care – from risk assessment to treatment – has been integrated into four U.S. primary care settings. Primary care adoption of LTBI services is critical for meeting U.S. TB elimination goals, given that primary care practitioners care for a larger proportion of the general population compared to public health clinics (8,9,11,21). By embedding routine risk assessments, IGRA testing over TST, and prescription of short-course, rifamycin-based treatments into the primary care workflow, these sites demonstrate that LTBI care can be feasibly decentralized from public health TB programs.

Each health system in this study served a large population at higher risk of TB infection, specifically those who are non-U.S.–born, a population disproportionately affected by TB in the United States (12). Average test positivity was in alignment with estimates for comparable populations at increased risk for TB (16%; range 10–25%) (3,19,22,23), and contributes to the growing body of literature measuring LTBI care outcomes in comparable settings (1,3,10,24). Compared to IGRA usage (37%) across 15 public health settings participating in TBESC-II (2011–2021) (25), our study sites demonstrated a higher adoption of IGRA testing (89%; range: 61–100%). Further, this high use of IGRA testing contributes evidence of the continued shift from TST to IGRA use following CDC guidelines for using QuantiFERON-TB (QFT) in 2003 (26), QFT-Gold in 2005 (27), and QFT-Gold In-Tube and T-Spot in 2010 (28,29).

Our analysis demonstrated that most LTBI workflows depend largely on the discretion of the PCP. Only Site D routinely supported PCPs by task-sharing LTBI care management with medical assistants who separately handled administrative, TST-based testing (Supplemental figures). Although Site A often referred patients diagnosed with LTBI to public health TB clinics for treatment, PCPs could still access patients’ visit information, potentially reducing losses to follow-up. Among the two sites with PCP-facing EMR tools for LTBI care management, the effect on physicians was unmeasured (Sites A and B), so the impact and uptake of these tools are unknown. This variability aligns with broader challenges described in the literature, including EMR alert fatigue for healthcare practitioners, competing health priorities for the patient, and lack of system-level TB champions (15,23). While EMR-based tools can support TB screening, their success depends heavily on practitioner engagement, EMR design usability, and integration into existing workflows (9,23).

Inconsistency in demographic data collection, particularly country of birth, in EMR hampers identification of individuals at risk for TB based on key epidemiologic factors. Routine and structured demographic documentation could improve screening precision and coverage (8,24,30–34). However, the collection and quality assurance of variables such as country of birth may be onerous and depends on dedicated resources, such as patient registration specialists and EMR technical support available to the health system.

Although all study sites implemented CDC-recommended practices for LTBI screening and care management, their workflows differed in both approach and consistency. Paired with PCP support strategies, including EMR prompt optimization, trainings, and task-shifting, primary care systems could reduce burden on the physician and improve consistency in TB screening and care management over time. Prior studies have documented several barriers to the provision of LTBI care in similar or community health care systems, including TBESC-II Part C (2018–2022) (35) where in-depth provider interviews revealed the following challenges and barriers to improving LTBI care: lack of knowledge of risk factors for LTBI, assumptions used for country of birth (i.e., limitations of using language as a proxy), lack of a standardized system for tracking patients prescribed LTBI medication, and provider concern regarding testing and treating persons with comorbidities (e.g., hepatitis B virus infection). Some of these challenges manifest as variability within workflows, suggesting systemic improvements like training staff in current LTBI screening and care guidelines, shifting tasks from PCPs to other medical staff, automating EMR processes to reduce care gaps and loss-to-follow-up, and adoption of culturally and linguistically appropriate outreach and patient education could improve could strengthen LTBI care management.

This study is limited in scope to primary care clinics serving at least 10,000 non-U.S.–born persons annually and was not designed to evaluate LTBI care outcomes or assess implementation effectiveness. The information presented here is based on self-reported workflows and site-level summaries of protocols and other documentation that may not reflect practices across all PCPs in each health system. The sites received funding to improve LTBI testing and treatment and therefore represent health systems actively invested in improving TB and LTBI care. Thus, our findings may not be representative of primary care across the United States or in regions substantially different than the ones described here. Finally, this study is limited to reporting on LTBI testing metrics; EMR-extracted information on LTBI diagnosis and treatment outcomes were not available for this broader population that encompassed all primary care patients. Despite these limitations, our findings still offer valuable, real-world insight for implementing LTBI care into primary care.

Reactivation of LTBI remains the primary driver for TB incidence in the United States (1,2). Therefore, TB prevention in primary care clinics is pivotal in identifying and treating infection before disease develops (13,14). Documenting how LTBI care has been incorporated into four primary care systems may inform practical strategies for TB prevention. These real-world insights may support other primary care systems serving populations at risk for TB as they consider implementation of LTBI care.

## Data Availability

All data produced in the present work are contained in the manuscript

