## Supplemental Figures for "Latent tuberculosis infection workflows in four primary healthcare systems in the United States"

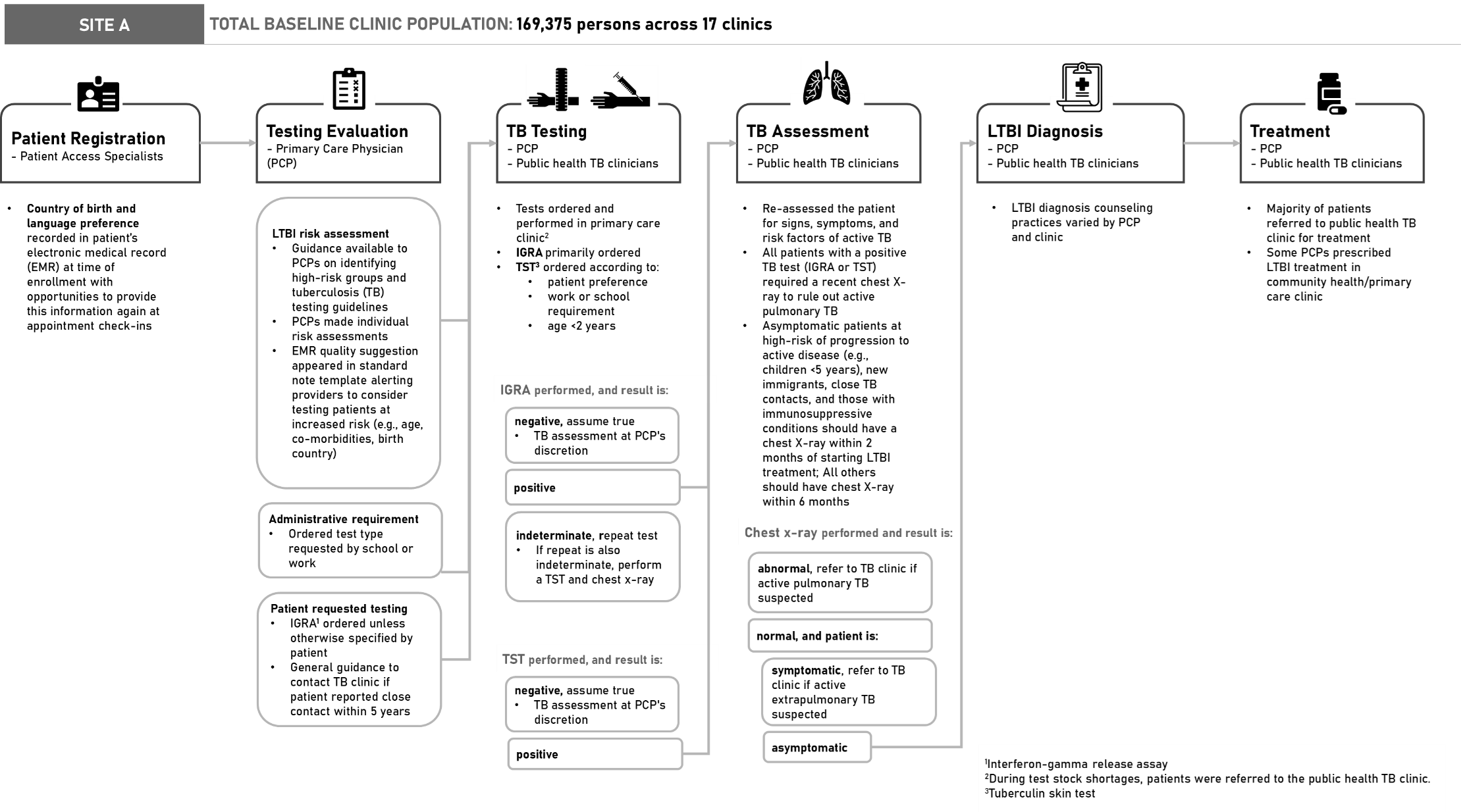

**Supplemental Figure 1.** Summary of the latent tuberculosis infection (LTBI) care workflow and clinical practices at Site A, a primary healthcare system participating in the Tuberculosis Epidemiologic Studies Consortium-III, 2020–2022.

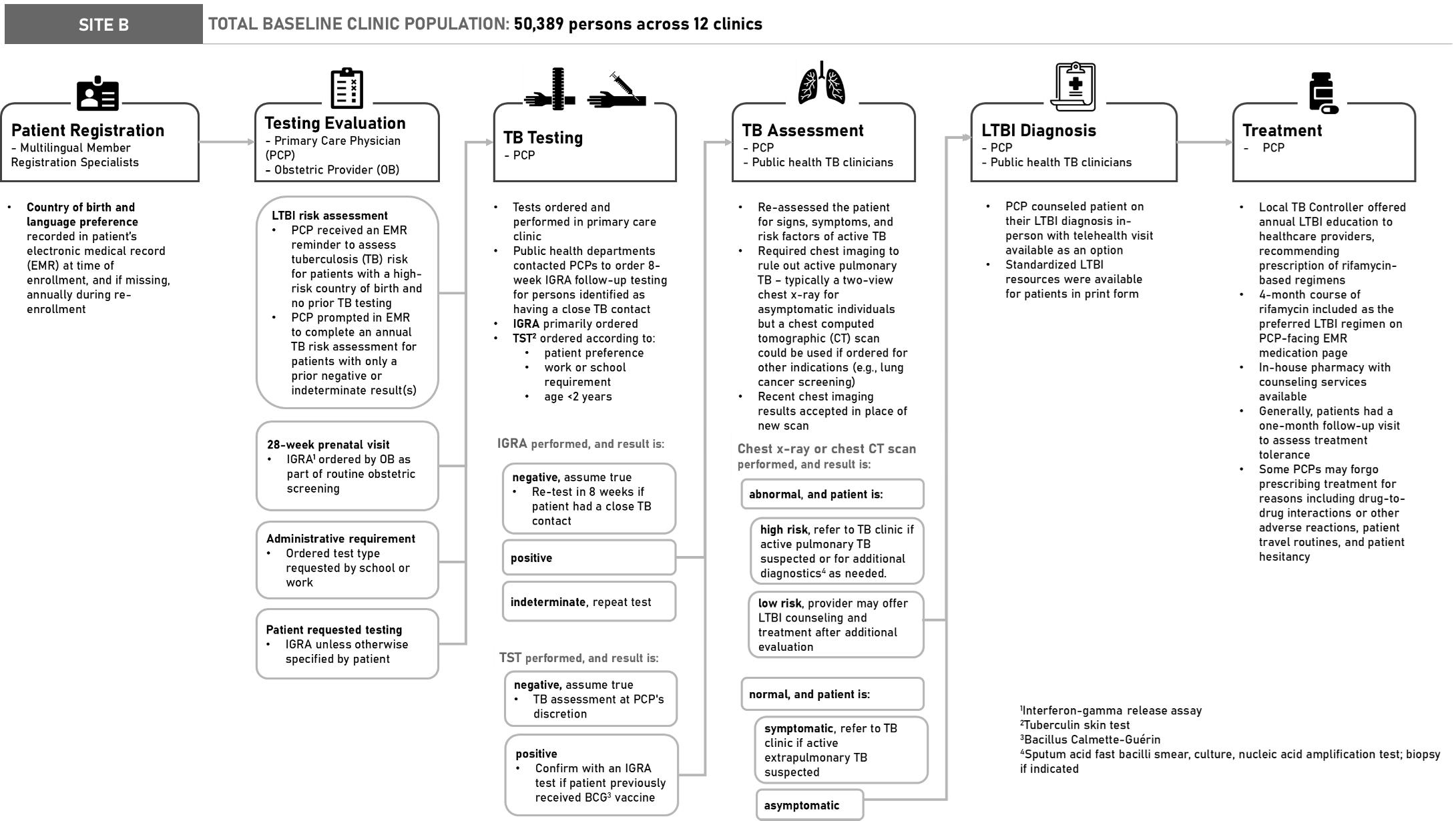

**Supplemental Figure 2.** Summary of the latent tuberculosis infection (LTBI) care workflow and clinical practices at Site B, a primary healthcare system participating in the Tuberculosis Epidemiologic Studies Consortium-III, 2020–2022.

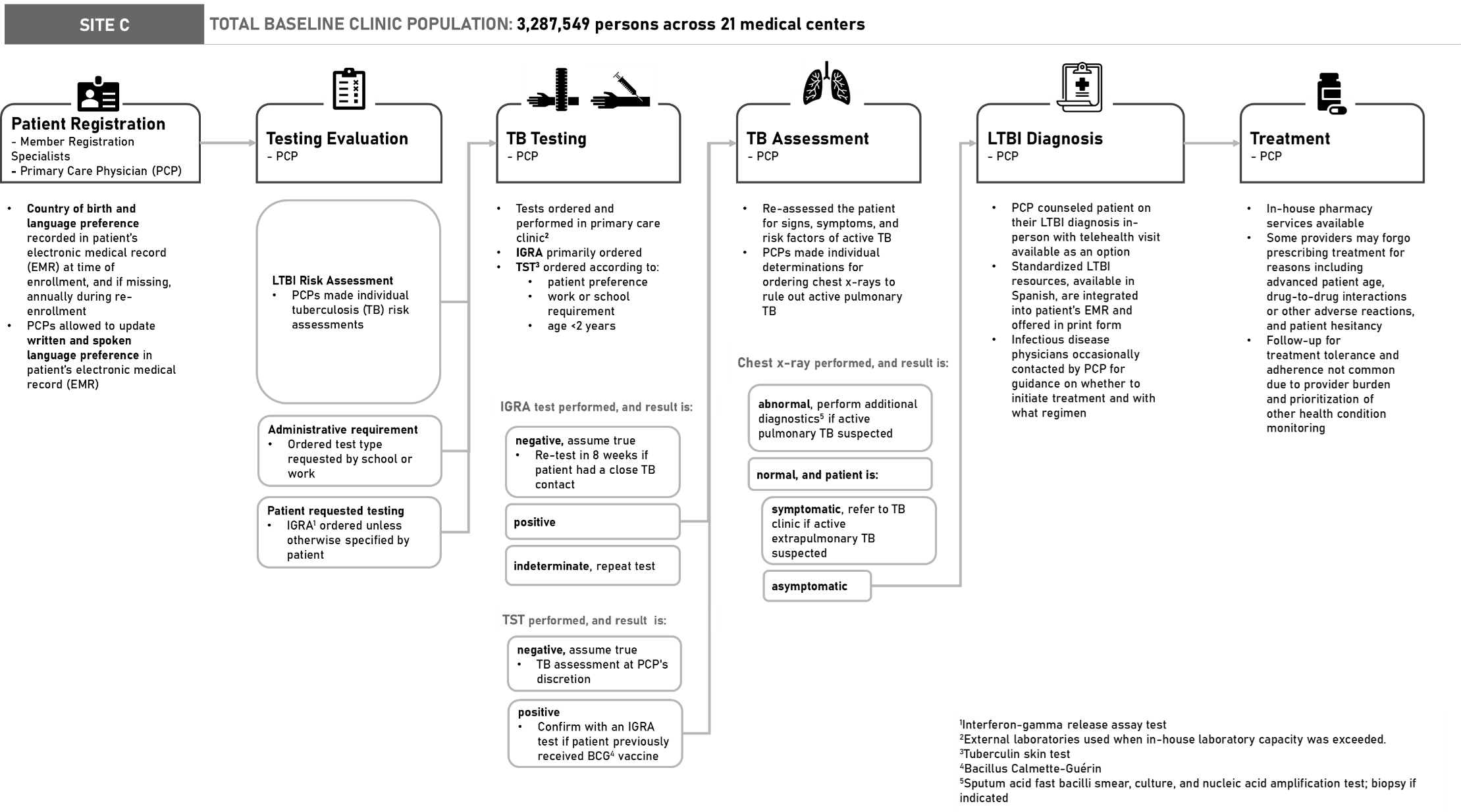

**Supplemental Figure 3.** Summary of the latent tuberculosis infection (LTBI) care workflow and clinical practices at Site C, a primary healthcare system participating in the Tuberculosis Epidemiologic Studies Consortium-III, 2020–2022.

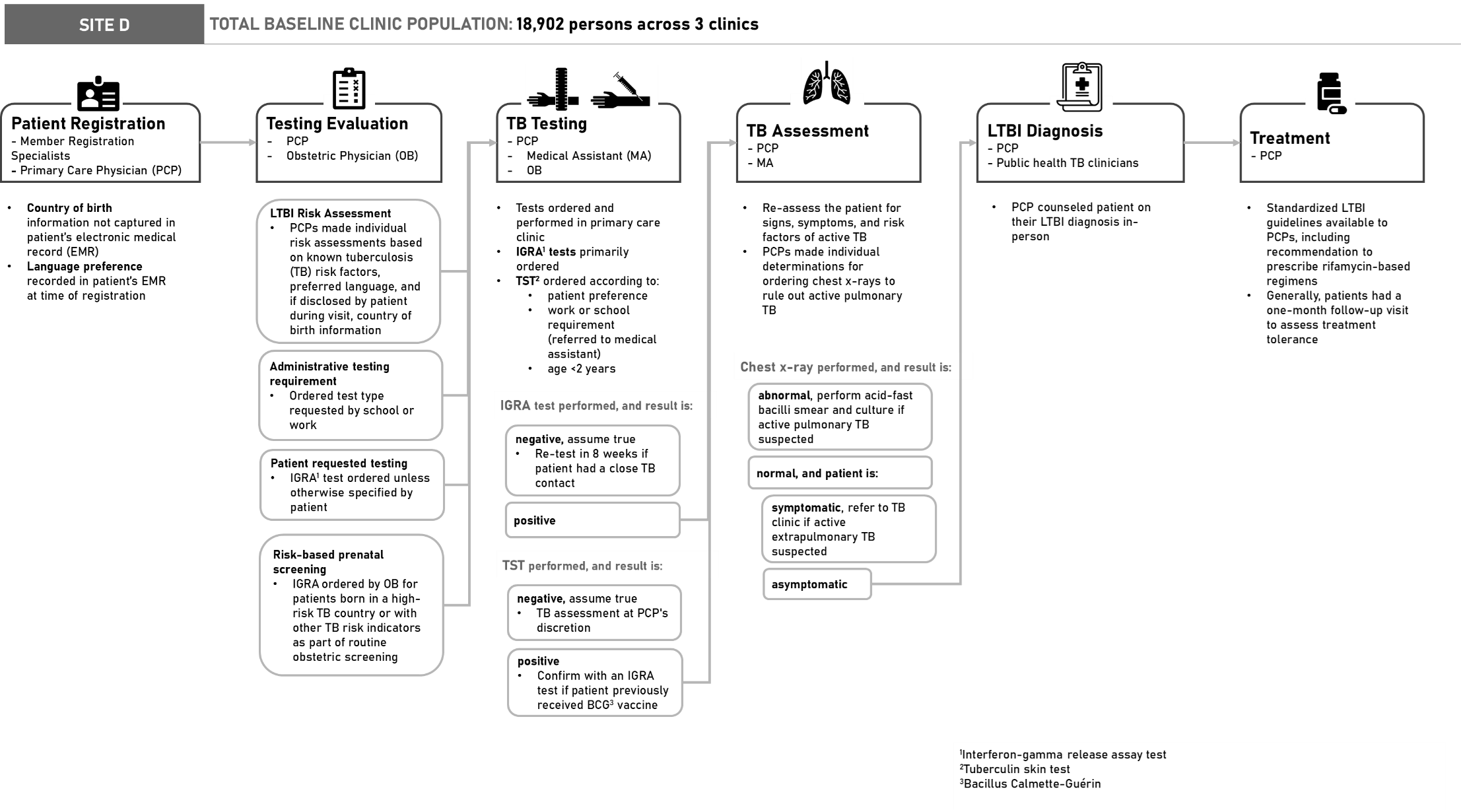

**Supplemental Figure 4** Summary of the latent tuberculosis infection (LTBI) care workflow and clinical practices at Site D, a primary healthcare system participating in the Tuberculosis Epidemiologic Studies Consortium-III, 2020–2022.
